# Explainable machine learning for real-time hypoglycaemia and hyperglycaemia prediction and personalised control recommendations

**DOI:** 10.1101/2022.03.23.22272701

**Authors:** Christopher Duckworth, Matthew J Guy, Anitha Kumaran, Aisling Ann O’Kane, Amid Ayobi, Adriane Chapman, Paul Marshall, Michael Boniface

## Abstract

**Background:** The occurrences of acute complications arising from hypoglycaemia and hyperglycaemia peak as young adults with type 1 diabetes (T1D) take control of their own care. Continuous glucose monitoring (CGM) devices provide real-time blood glucose readings enabling users to manage their control pro-actively. Machine learning algorithms can use CGM data to make ahead-of-time risk predictions and provide insight into an individual’s longer-term control.

**Methods:** We introduce explainable machine learning to make predictions of hypoglycaemia (<70mg/dL) and hyperglycaemia (>270mg/dL) 60 minutes ahead-of-time. We train our models using CGM data from 153 people living with T1D in the CITY survey totalling over 28000 days of usage, which we summarise into (short-term, medium-term, and long-term) blood glucose features along with demographic information. We use machine learning explanations (SHAP) to identify which features have been most important in predicting risk per user.

**Results:** Machine learning models (XGBoost) show excellent performance at predicting hypoglycaemia (AUROC: 0.998) and hyperglycaemia (AUROC: 0.989) in comparison to a baseline heuristic and logistic regression model.

**Conclusions:** Maximising model performance for blood glucose risk prediction and management is crucial to reduce the burden of alarm-fatigue on CGM users. Machine learning enables more precise and timely predictions in comparison to baseline models. SHAP helps identify what about a CGM user’s blood glucose control has led to predictions of risk which can be used to reduce their long-term risk of complications.

## Introduction

People with type-1 diabetes (T1D) face a daily balance to keep their blood glucose levels within safe levels (i.e. ‘in-range’). Severe complications are prevalent and arise from glycaemic variability, low blood sugars (hypoglycaemia) and high blood sugars (hyperglycaemia)[1]. For hypoglycaemic incidents alone, the requirement for emergency assistance may be as high as 7.1% per year [2] and could account for 6-10% of deaths for those with T1D [3, 4]. Long-term impacts of hypoglycaemia include impacts on cognition and potential links with dementia[5]. In addition, frequent hyperglycaemia can lead to short-term risk such as diabetic ketoacidosis and long-term complications such as retinopathy, neuropathy, nephropathy, and cardiovascular disease[6-8]. Effective glucose management for adolescents and young adults living with T1D is challenging[9, 10], due to the multiple transitions taking place in their lives, including puberty, relationships, the move to more independent living and diabetes self-care, and also the transfer from paediatric to adult clinical care teams. Parental fear of severe complications is prevalent throughout these transitional years[11-13].

Continuous glucose monitoring (CGM) enables regular automated readings of estimated blood glucose levels, providing immediate insight into blood glucose control. CGM has been demonstrated to reduce the risk of both hypoglycaemia and hyperglycaemia, along with reducing daily glycaemic variability for users with type-1 diabetes[14-16]. In addition to mitigating short-term risk of severe hypoglycaemia and hyperglycaemia, compliance of wearing CGM devices has been shown to improve glycosylated haemoglobin HbA1c levels, which, if sustained, reduce long-term complication risks[17, 18]. The magnitude of reduction in HbA1c from CGM usage is dependent on the user’s original HbA1c value; i.e. those at highest risk of complications from poorer control are likely to benefit the most [16]. Specific to young adults, Laffel et al. [19] demonstrate a clear improvement in HbA1c for those utilising CGM.

Real-time CGM devices provide alerts for users when their blood glucose falls above or below a desired range. T1D management can be aided further by having *ahead-of-time* predictions so individuals can identify risk early and better plan self-care activities, such as insulin dosages. Simple threshold-based algorithms have been able to successfully predict hypoglycaemia 30 minutes in advance (e.g. Medtronic-640 ‘SmartGuard’[20]). More complex statistical models and machine learning algorithms enable more accurate prediction and are able to extend this prediction horizon[21-28]. Dave et al. [23] emphasize the importance of feature extraction when generating predictions of hypoglycaemia in CGM data. Generating features that are both predictive in models and insightful for understanding a user’s blood glucose control is a difficult balance.

In this work, we make two novel contributions: algorithms tailored to young adults and explanations. First, we introduce machine learning models to predict hypoglycaemia (<70mg/dL) and hyperglycaemia (>270mg/dL)[29] with a trustworthy 60-minute prediction horizon for young adult users of CGM. While CGM risk prediction is a well explored topic, more must be done to understand what led to increased risk for an individual so they can be proactive. We introduce using *explainable* machine learning, to not only predict risk, but to automatically identify the most important factors in an individual’s CGM data that led to increased risk. Explanations have no detrimental impact on model performance. We provide a framework in which machine learning can be used to:

1. Provide real-time predictions of hypoglycaemia and hyperglycaemia (Results - Model Evaluation) using intuitive features (Methods – Features) generated from CGM data (Methods – Data).
2. Automatically identify the most important features that have led to predictions of risk for each CGM user over a given time-period (Results – Model Explanation).
3. Provide personalised control recommendations for each CGM user to help with their T1D management (Results – User Interface).

## Methods

### Data

We make use of publicly available data from “A Randomized Clinical Trial to Assess the Efficacy and Safety of Continuous Glucose Monitoring in Young Adults 14-<25 with Type 1 Diabetes” (CITY)[19]. By design, the study recruited adolescents and young adults with T1D (duration > 12 months) exhibiting poorer glycaemic control (HbA1c 7.5-<11.0%), most likely to benefit from CGM usage [16]. Study participants were randomly assigned to either CGM (Dexcom G5) or regular blood glucose meter (finger-prick) monitoring. The CGM users were compared to the control group using HbA1c levels after six months of usage. After six months, all study participants were provided with CGM devices and HbA1c tracked for a further six months.

We make use of CGM data from 153 people living with T1D in the CITY study, where users were provided CGM devices for 6-12 months; totalling over 28,000 days of usage data. In addition to CGM data, basic screening information and the most recently recorded HbA1c test result were used to generate predictions.

### Features

To utilise CGM data for hypoglycaemic and hyperglycaemic prediction, we generate a total of 30 features which summarise a young adult’s CGM data on different timescales. Blood glucose control is summarised on short-term (one hour), medium-term (one day) and long-term (one week) baselines prior to the current CGM reading. This is combined with six features that characterise basic patient information. A complete description of all generated features are given in Table 1. Features are generated at the point of each unique CGM reading. Features are only used in modelling if the CGM device has been used for >=80% for the prior week.

**Table 1:**
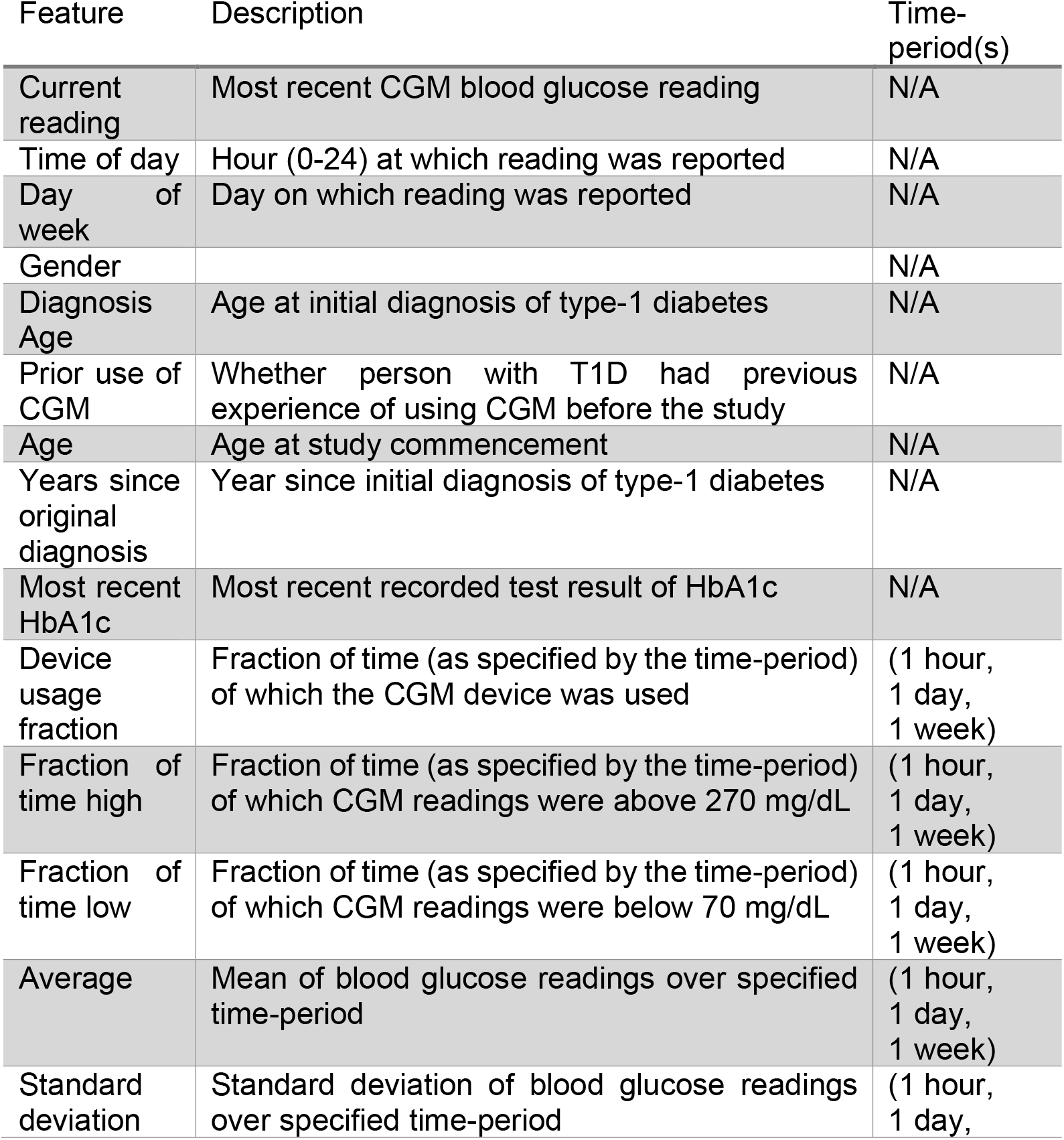
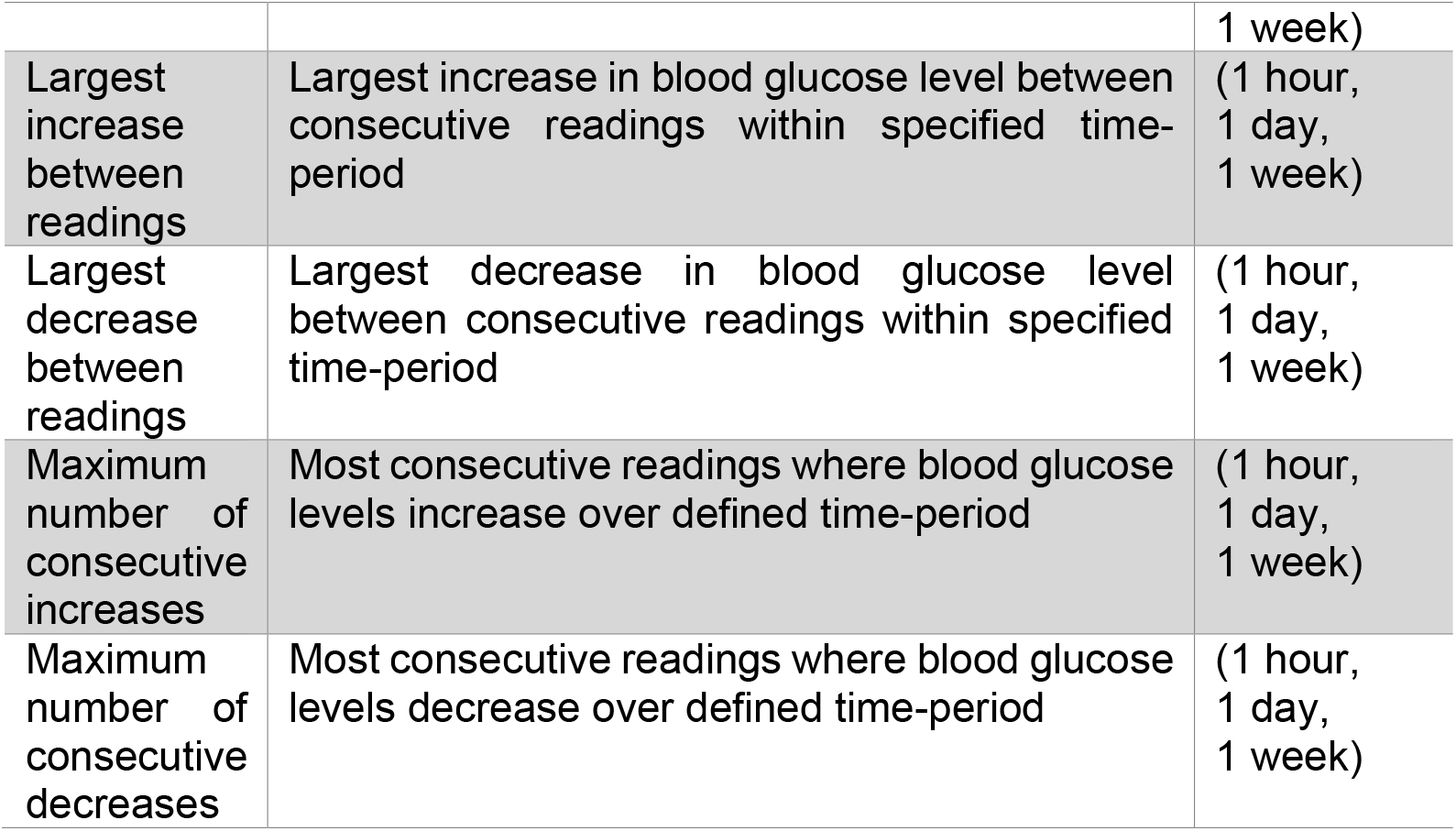
Summary of input features used by the models to make predictions. A sub-set of features are computed for various time-ranges (i.e. 1 hour, 1 day, 1 week) and considered as independent features.

### Targets

To generate targets for our model predictions, we generate two binary variables referring to hypoglycaemic (< 70 mg/dL) and hyperglycaemic (> 270 mg/dL) events. A feature set is generated for each unique CGM reading, at which point we check if the CGM user’s blood glucose level falls within these regions in the following 60-minutes (i.e. positive prediction). Blood glucose readings already within the hypoglycaemic or hyperglycaemic regions are removed from the modelling dataset to avoid artificially boosting model performance metrics. Figure 1 shows a schematic of blood glucose levels through a given day, regions of hypoglycaemia and hyperglycaemia and timestamps of model predictions prior (i.e. target).

**Figure 1:**
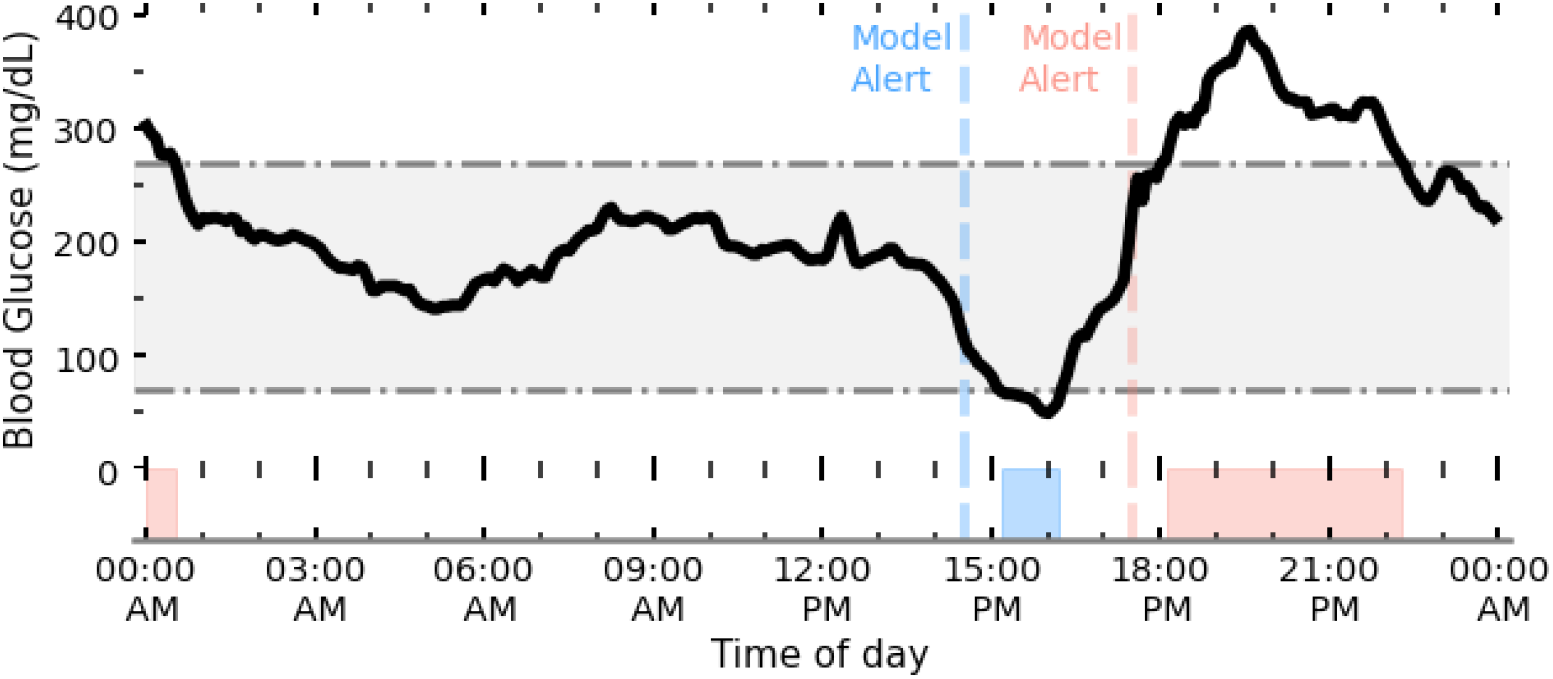
Schematic of blood glucose levels (black line) for a young adult with T1D tracked by CGM. The grey shaded region shows the desired range to keep blood glucose levels between (70mg/dL < BG < 270mg/dL). Our algorithm aims to predict (ahead-of-time) when a person with T1D will go below (hypoglycaemia) and above (hyperglycaemia) this range. Regions of low and high blood glucose are shaded blue and red respectively, with the corresponding first prediction event horizon (i.e. when our model first made a positive prediction of hypo/hyper) shown by the dashed line.

### Modelling

To determine the added value of machine learning we evaluate a baseline heuristic model, a logistic regression model and a gradient boosted tree-based model for both hypoglycaemia and hyperglycaemia prediction. Our baseline heuristic model is equivalent to a blood glucose threshold alert (i.e. predicting hypoglycaemia and hyperglycaemia within 60-minutes if blood glucose levels fall below 110mg/dL or go above 240mg/dL respectively). Our logistic regression model is aimed to emulate basic CGM alerts which extrapolate linear trends along with thresholds to make hypoglycaemia or hyperglycaemia predictions.

Finally, we make use of the XGBoost framework to implement a tree-based machine learning algorithm [30]. XGBoost makes use of an ensemble of weak learners (i.e. small trees) that are trained stage-wise through gradient boosting. This reduces overfitting while preserving or lowering variance in the prediction error [31], which frequently leads to gradient boosted trees outperforming other tree-based methods. Additionally, XGBoost naturally deals with continuous, binary/discrete, and missing data consistently; all of which are represented in our dataset. Model hyperparameters for our XGBoost models were selected using five-fold cross-validation of the complete training set using a sampler (Tree-structured Parzen Estimator) implemented with the Optuna library[32].

We randomly separate our CGM data into a hold-out test set (25%) and a training set (75%). Our supervised models (i.e. logistic regression and XGBoost) learn from the training set, and all models are evaluated using the same test sample. Overall, model performance was evaluated using the Area Under the Receiver Operating Curve (AUROC) and average precision, along with fixed measures of specificity and sensitivity.

### Model explanability

Historically, machine learning algorithms are considered ‘black-boxes’ with little understanding of how predictions have been made. However, recent advances in *explanability* have led to individual predictions of tree-based algorithms being readily explainable[33].

To attribute the relative importance of each feature in predicting both hypoglycaemia and hyperglycaemia risk for our XGBoost model, we make use of the TreeExplainer algorithm as implemented in the SHAP (SHapley Additive exPlanations) library[33-35]. TreeExplainer efficiently calculates Shapley (SHAP) values[36], which aim to attribute payout (i.e. the prize) between coalitional players of a game. In the context of machine learning, SHAP values amount to the marginal contribution (i.e. change to the model prediction) of a feature amongst all possible coalitions (i.e. combinations of features). Practically, this means that for every individual prediction (negative or positive), the relative importance of every feature can be evaluated.

There is a rich history of global interpretation for machine learning models which summarise the average overall importance of features on predictions as a whole[37]. In a medical setting, however, tailored explanations for individuals are paramount, maximising the ability to understand their own data and ensure every person is evaluated fairly[38]. Shapley values are *locally accurate*, meaning that they can explain which features were relatively most important for an individual prediction (i.e. a hypoglycaemic or hyperglycaemic event). In addition, Shapley values are consistent (the values add up to the actual prediction of the model) meaning they can also be used to check the global importance of a feature. Feature importance can therefore be checked periodically by averaging over a fixed time-period. Practically, this means that for a CGM user over a given time-period, the most important features leading to a prediction of hypoglycaemia or hyperglycaemia can be automatically evaluated. This gives immediate insight about an individual’s blood glucose control, and intuition about what may be increasing their risk. Presenting reliable predictions with intuitive explanations, would enable users to be proactive in their control. Insightful control recommendations could empower users to feel closer to being on ‘auto-pilot’ (i.e. minimising the cognitive load burden).

We choose to implement SHAP over other local explainer algorithms (e.g. Lime[39]) since SHAP offers mathematical guarantees of trustworthiness (local accuracy, missingness, and consistency) which adhere to strict medical governance guidelines[33], and offers consistency between local explanations meaning global importance can be computed as well.

## Results

### Model evaluation

In Figure 2, we compare the performance of our baseline heuristic model against the machine learning classifiers (i.e. logistic regression and XGBoost). Performance is evaluated by the AUROC characteristic by comparing the model predictions of hypoglycaemia (left) or hyperglycaemia (right) 60-minutes ahead-of-time to the actual future readings. For hypoglycaemia, the baseline model achieved an AUROC of 0.811, the logistic regression 0.930 (95% CI:0.929-0.931) and XGBoost 0.998 (95% CI:0.998-0.998) evaluated on our hold-out test set. All confidence intervals (CI) are estimated from bootstrapping (sampling with replacement) for 500 resamples per model.

**Figure 2:**
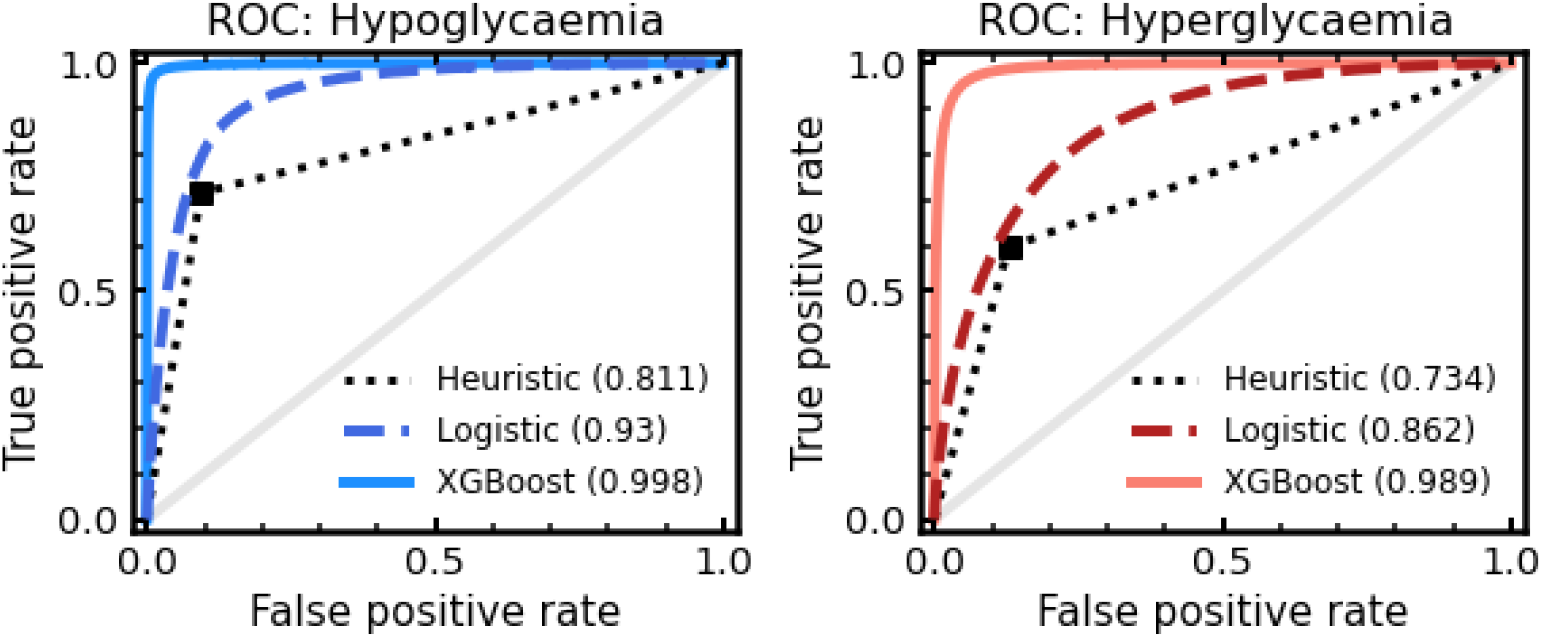
Receiving operator characteristic (ROC) for our models of hypoglycaemia (left) and hyperglycaemia (right) prediction. In each panel, a XGBoost model (solid line) and a logistic regression model (dashed line) are compared to a baseline heuristic (dotted line). A zero skill model is represented by the solid grey line. The total area under each curve (i.e. AUROC score) is given in the brackets.

Both machine learning models demonstrated excellent predictive power for hypoglycaemia, with a clear advantage in using XGBoost. We note that despite its crudeness, our baseline heuristic model also performs well; demonstrating the use of threshold-based alerts on CGM devices in forward planning. Regardless, a more powerful predictive model means a lower false-alarm rate can be achieved, while maintaining the safety of the predictions. Reducing alarm-fatigue for CGM users is an important goal, and more skilful models help enable this. In Table 2, additional measures of model skill are given, including average precision, sensitivity, and specificity. Sensitivity and specificity are evaluated from dichotomising model predictions at probability P=0.5. Again, we find a clear performance increase for our XGBoost model, in-keeping with the high performance of decision tree based methods[40] and commercial hybrid loop systems[41].

**Table 2:** Summary of model performance metrics for both hypoglycaemia and hyperglycaemia prediction. A baseline heuristic, logistic regression and an XGBoost model are evaluated for each target. Summary statistics (AUROC and average precision) are shown with 95% CI in square brackets Sensitivity and specificity are evaluated from dichotomising model predictions at probability P= 0.5.

| MODEL | AUROC | AVERAGE PRECISION | SPECIFICITY (P<sub>THRES</sub> = 0.5) | SENSITIVITY (P<sub>THRES</sub> = 0.5) |
| --- | --- | --- | --- | --- |
| **Hypoglycaemia** |  |  |  |  |
| Heuristic | 0.811 | 0.121 | 0.906 | 0.716 |
| Logistic Reg. | 0.930 [0.929-0.931] | 0.244 [0.240-0.247] | 0.827 | 0.905 |
| XGBoost | 0.998 [0.998-0.998] | 0.953 [0.951-0.954] | 0.994 | 0.945 |
| **Hyperglycaemia** |  |  |  |  |
| Heuristic | 0.733 | 0.258 | 0.872 | 0.595 |
| Logistic Reg. | 0.862 [0.861-0.862] | 0.453 [0.450-0.456] | 0.752 | 0.817 |
| XGBoost | 0.989 [0.989-0.990] | 0.931 [0.930-0.932] | 0.931 | 0.970 |

High performance is also seen for hyperglycaemia, with the baseline model achieving an AUROC of 0.734, the logistic regression 0.862 (95% CI:0.861-0.862) and XGBoost 0.989 (95% CI:0.989-0.990). Average precision, sensitivity, and specificity demonstrate similar trends with XGBoost being the most skilful. For each modelling approach we note that the model skill is lower for hyperglycaemia prediction in comparison to hypoglycaemia, suggesting prediction of lower blood glucose events is better suited to our modelling choices.

### Model explanation

In addition to increased predictive power, the added value of machine learning models can be demonstrated through explanations. Using SHAP we can evaluate the relative importance of features for a given positive prediction of hypoglycaemia or hyperglycaemia. SHAP is applied post model construction and therefore has no negative implications for performance. Figure 3 shows the overall relative importance of every input feature for predicting hypoglycaemic (left-panel) and hyperglycaemic (right-panel) events. The relative importance of a feature is quantified by the absolute average SHAP value. Since SHAP values are consistent across predictions, they can be averaged for individual CGM users, across any time-range, to provide immediate insight.

**Figure 3:**
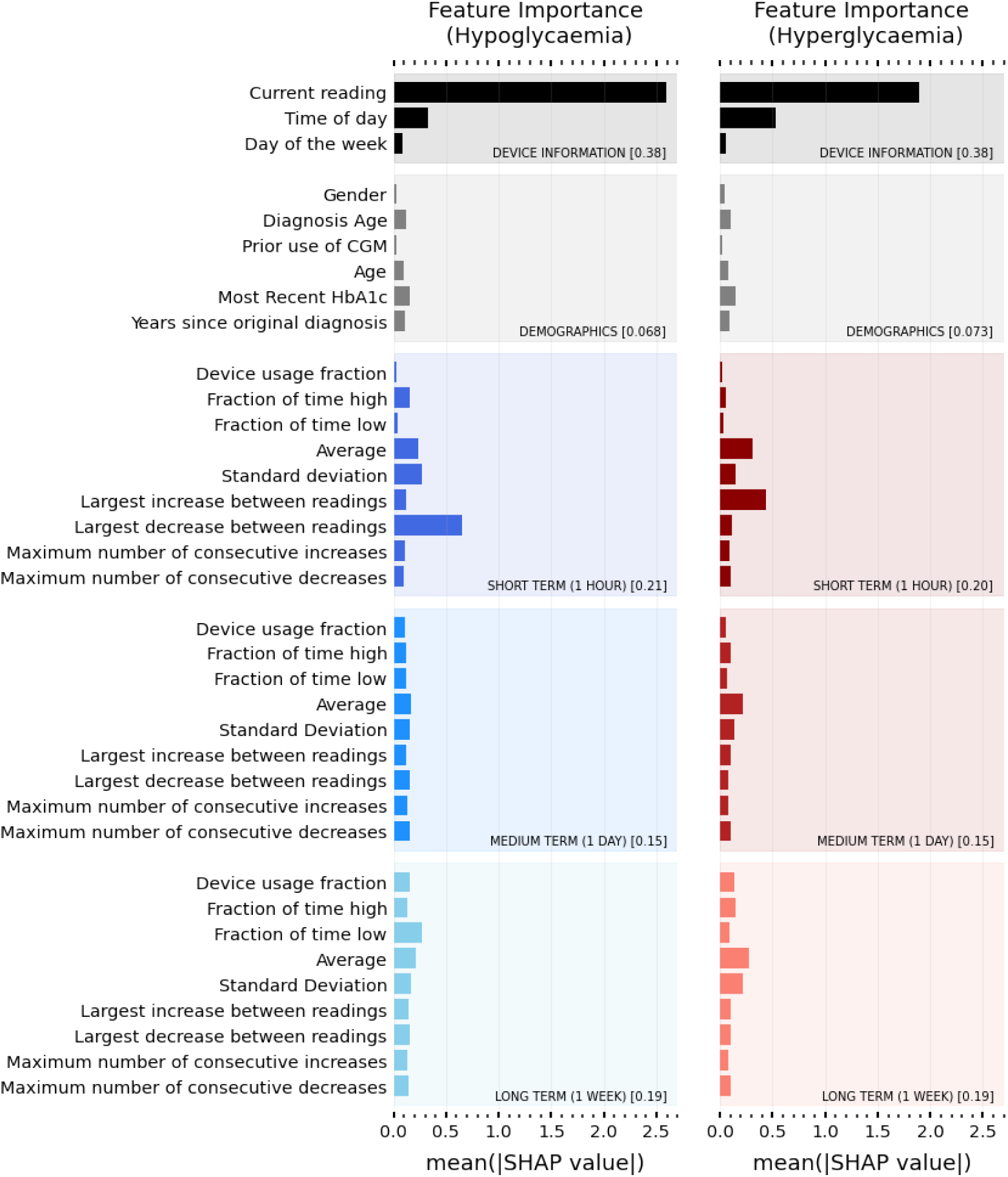
Overall importance ranking of input features for predicting hypo (left panel) and hyper (right panel) risk. Average (absolute) SHAP value for predictive features over all study participants. A higher value corresponds to a more important feature in decision making. Features are grouped into categories (Device information, Demographics, Short term (1 hour), Medium Term (1 day), Long term (1 week)). The fractional contribution (i.e. sum over all features in that category) of a given category is given in the square brackets.

Here we provide the average relative importance for all CGM users in the study, but this diagram is trivially made for individual users. Unsurprisingly, the user’s current blood glucose reading is most important for the model to make predictions of both hypoglycaemia and hyperglycaemia. Time of day is also important, providing insight into the sleep and eating, physical activity and stress level habits of the CGM user and their relationship with blood glucose. Sudden drops (or increases) in blood glucose are important for predicting hypoglycaemia (hyperglycaemia) as shown by the short-term largest decrease (increase) between readings. Interestingly the long-term fraction of time low is found to be reasonably predictive of hypoglycaemic events, providing immediate insight into certain user’s control habits.

### User interface

Despite CGM providing a wealth of information to both users and clinicians, the sheer volume of data makes it hard to quickly draw conclusions about blood glucose control. Quick summary metrics such as the fraction of time-in-range (e.g. 70mg/dL<BG<270mg/dL) are the baseline for assessing control. By considering the most predictive model features that led to predictions of hypoglycaemic or hyperglycaemic events, we can draw further personalised insights into an individual’s blood glucose control. In Figure 4, we present a prototype dashboard which summarises a randomly selected user’s CGM data over a given month, along with potential insights derived from explainable machine learning. In addition to metrics such as time above or below range, we provide the user’s average blood glucose through the day, along with the most likely times for our model to predict hypoglycaemia (red, above green line) or hyperglycaemia (blue, below green line) for the individual. We select the top features for predicting both hypoglycaemia and hyperglycaemia for the user and summarise this information as control recommendations in the grey box. This provides a quick glance into the specifics of the user’s blood glucose control; enabling the user to be better informed to avoid potential events in the future. One AI insight (grey box) for this user is that they tend to go high at specific times of day. Looking at the fraction of time spent high on the dashboard through the day (red box and histogram), this peaks around 21:00pm, hence the user should consider insulin dosages around their evening meal.

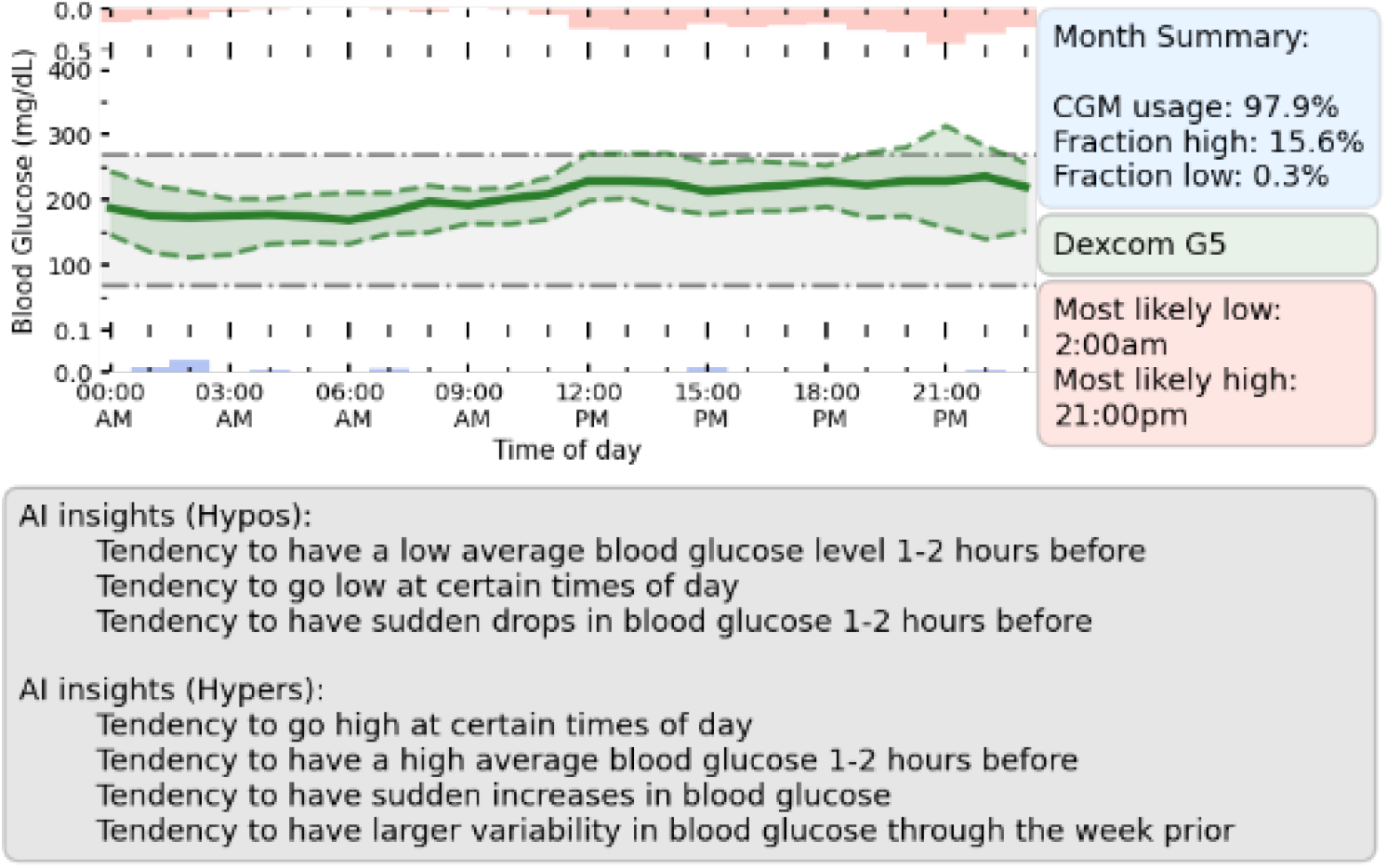

## Discussion

The key contributions of our work are as follows:

1. Machine learning models with state-of-the-art performance for predicting hypoglycaemia (AUROC:0.998) and hyperglycaemia (AUROC: 0.989) 60-minutes in advance. This performance is high relative to simple algorithms[42-44] and comparable machine learning approaches[23, 45].
2. With careful feature engineering, we have demonstrated how machine learning explanations (SHAP) can be utilised to understand specifics about an individual’s control. SHAP also adds transparency to model predictions, aiding assurance that all individuals are evaluated fairly.
3. Provided a prototype dashboard to help young adults with T1D and clinicians make use of CGM data and the insight from machine learning explanations.

Technological advances represent a significant opportunity to help reduce self-care burden on an individual with T1D, and reduce the risk of health complications arising from poor glycaemic control. In particular, for young adults, automated feedback from CGM may be an important tool for reducing risk, at times of transition (from paediatric to adult care units) and where glycaemic control can be at a minimum.

Ahead-of-time machine learning predictions are of personal and clinical value as they give the CGM user more time to adjust self-care and reduce risk. Our tree-based model demonstrated a significant performance increase relative to threshold based and linear models. This performance increase is vital for reducing alert burden on the user, since more certain predictions require less total alerts while maintaining safety of the device.

Despite the wealth of information provided by CGM devices, part of the problem is deriving quick insight that is useful for people with T1D, their family carers, and clinicians[46, 47]. Machine learning explanations can help summarise what specifics in an individual’s glycaemic control led to increased risk of either hypoglycaemia or hyperglycaemia. Used in combination with directly derived metrics (e.g. time-in-range), their utility can be in providing quick-glance specific recommendations about how to reduce risk.

### Limitations

Limitations of this work include the reliance on the user to comply in using the CGM device. For our results, we only generate predictions when the user has used the device for 80% of the prior week. While predictions can still be generated with a lower usage compliance, this will inevitably decrease prediction performance, and care must be taken about when machine learning enhancement can be implemented safely. Furthermore, while current CGM devices are generally accurate, they are not infallible and considerations must be made for the safety of systems reliant on their accuracy[48].

Another limitation of this study is the lack of insulin and carbohydrate data. Including this information could enable specific recommendations about insulin and carbohydrate dosages through the day. Including information tracked by smart watches, such as physical activity and stress levels, would not only improve predictions, but provide far more powerful intuitive recommendations. Having contextual information (e.g. high stress levels or even self-reported event markers such as drinking, sickness or exercise) would be critical for empathetic recommendations and reducing burden for the user.

In this work we chose to train hypoglycaemia and hyperglycaemia models using data from all CGM users in our cohort. In practice it may be more suitable to train *individual* models per CGM user, which may be better tailored to the individual. However, it would be more complex to make direct comparisons between relative feature importance for different CGM users, and hence left outside the scope of this paper.

## Conclusion

We introduced a framework for high-performance prediction and explanation of hypoglycaemia and hyperglycaemia for young adults. Careful feature selection enables both accurate short-term risk prediction, and intuitive feedback about an individual’s blood glucose control. The key benefit of adopting a machine learning framework lies in the ability to provide more accurate ahead-of-time predictions (in comparison to more simplistic derived alerts), potentially reducing burden on the young adult potentially going through transition with their care practices. Combining these models with explanations enables both users and clinicians to gain immediate insight into an individual’s blood glucose control, automatically highlighting what specific trends lead to increased risk.

## Data Availability

Data is open source and can be found at https://clinicaltrials.gov/ct2/show/NCT03263494

## Acknowledgements

We acknowledge funding from UKRI Trustworthy Autonomous Systems Hub (Grant code: RITM0372366).

## Abbreviations

T1D: Type-1 Diabetes
CGM: Continuous Glucose Monitor
AUROC: Area Under the Receiver Operating Curve
SHAP: SHapley Additive exPlanations

